# A 10-Year Comparison of Injury Patterns and Outcomes between Migrants and Residents in a high-income country setting

**DOI:** 10.1101/2025.10.23.25338532

**Authors:** Xinyi Lin, Juin Bin Lee, Hwee Pin Phua, Beth Shin Rei Lau, Natasha Shi Hui Wee, Daniel Yong Jing Quek, Li-Tserng Teo, Raj Kumar Menon, Wei-Yen Lim, Hong Yee Lo

## Abstract

**Introduction:** Migrants are drawn to high-income countries (HICs) for employment, often in sectors deemed difficult, risky, and low paying. However, they experience more injuries and deaths and face barriers to healthcare. The World Health Organization has highlighted migrant health as a priority. While Hargreaves et al. and others studied migrant occupational health, most did not compare with residents. Lau et al. and Pega et al. did, but with limitations: the former focused on deaths, the latter judged the evidence to be poor. Few large studies in HICs directly compare injury outcomes and barriers between migrants and residents. Singapore, with a sizeable migrant workforce in construction, marine, and process industries, provides a suitable setting. We hypothesised that migrants sustain more work- and commute-related injuries, and poorer outcomes.

**Methods:** We performed a 10-year retrospective analysis (2013–2022) of injured patients at a central tertiary hospital in Singapore using its trauma registry. Migrants and residents were identified by registration numbers. Outcomes included injury severity, mortality, access barriers, and mechanisms of injury. Logistic regression adjusted for confounders.

**Results:** After excluding cases with missing data (31,555), 281,847 patients remained; migrants comprised 66,719 (23.7%). Migrants were predominantly young, male, and Chinese or Indian. Adjusted analyses showed migrants had lower odds of significant injuries (OR 0.87, 95%CI 0.81–0.93) and mortality (OR 0.64, 95%CI 0.48–0.84), but higher odds of discharging Against Medical Advice (OR 1.42, 95%CI 1.31– 1.55) and higher ED discharge prevalence (PR 1.04, 95%CI 1.04–1.05). Falls and vehicular accidents were leading mechanisms in both groups. Migrants had more falls from height (74.9%) and vehicular injuries (35.8%), while residents had more ground-level falls (46.0%).

**Conclusion:** Contrary to our hypothesis, migrants had lower odds of significant injuries and mortality. Protective mechanisms such as workplace safety policies may contribute. Yet, higher AMA and ED discharges suggest barriers to care. Findings highlight the need for prevention focused on falls from height and vehicular trauma, and continued efforts to reduce healthcare disparities in migrants.

## Background

Justice and medicine are intertwined disciplines, with healthcare disparities often revealing broader societal inequalities. An example is how migrants in both high-income and low-income countries deal with unmet needs for mental health, legal barriers in accessing healthcare, overuse of emergency services, underuse of primary healthcare services, as well as outright discrimination.^1,2^

The World Health Organisation (WHO) emphasised the need to prioritise research on health, migration, and displacement at the national, regional, and global levels^3^. This call reinforced existing works and encouraged further research. Hargreaves et al studied the prevalence of occupational health outcomes in migrants, but did not compare with resident populations.^4^ Similarly, other research on injury incidence, health services utility, injury types did not compare with resident populations.^5-7^ Lau et al did compare between migrants and local workers in a global systematic review, but focused narrowly on mortality.^8^ Pega et al also compared between migrants and non-migrants but assessed the quality of the evidence to be low to very low due to biases, indirectness, and imprecision.^9^ Other studies reported findings from low and middle income countries.^10-13^ Few large original studies offer high quality data for direct comparison of injury patterns and health outcomes between migrants and residents in a high income country (HIC) setting.

Singapore, a developed country with a population of 6.04 million, of which 1.86 million are migrants,^14^ offers an appropriate context for such research. Migrants make up a substantial proportion of the workforce, with 59% of migrants being semi-skilled and unskilled workers in the construction, manufacturing, and processes (CMP) industries,^15^ as well as domestic work, sectors that are typically shunned by locals. Additionally, migrants are not fully assimilated in their host countries. In Singapore, many live in purpose-built facilities with different transportation means, such as the use of motorcycles or are being transported on lorries.^16^

Like other high-income countries, Singapore has enacted legislations to protect and promote safety amongst migrants.^17^ While the living and work environments are different for migrants and residents, injuries that are deemed serious enough would be treated at the same hospital system, with a unified pre-hospital emergency care system. This provides the opportunity to investigate if disparities in migrant injury patterns and outcomes persist despite aspirations for equity of safety and healthcare in a HIC setting.

### Aims and Hypothesis

The aims of this study are to describe and compare the patterns and outcomes of injuries between migrants and residents.

The hypotheses of this study are, (1) migrants suffer poorer outcomes in terms of injury severity and mortality, (2) they face barriers when accessing healthcare, (3) and the mechanisms of injury are different between the two populations.

## Methods

### Study Design and Subjects

This study is a retrospective cohort study of injured patients from the trauma registry at Tan Tock Seng Hospital (TTSH) over a 10-year period, from January 2013 to December 2022. TTSH is a 1,500-bedded acute tertiary hospital within the NHG Health Group that serves the central and northern regions in Singapore, with a catchment of approximately 1.5 million people.^18^

The registry captures patients presented to the hospital with a trauma-related Systematised Nomenclature of Medicine Clinical Terms (SNOMED CT) or International Classification of Diseases (ICD) diagnosis.

Patients are classified into three tiers based on their Injury Severity Scores (ISS) which range from 1 to 75. Tier 1 patients (ISS ≥ 16) have severe injuries. Tier 2 patients (ISS 9 - 15) have moderate injuries, while Tier 3 patients (ISS ≤ 8) have minor injuries. For this study, moderate/ severe injuries are labelled as significant injuries.

### Data Collection

The data was collected and maintained by a team of dedicated trauma coordinators and undergoes regular auditing by the Ministry of Health. Patients younger than 21 years old or had missing data regarding their national identification number prefixes, age, or gender were excluded.

The variables included demographic data, residential status, injury severity, mechanisms of injury, disposition at the emergency department, and mortality during the healthcare encounter. Residency status was determined using the prefix alphabet of identification numbers. Residents have prefixes S or T. Migrants have prefixes G, F or M. Unknown patients are assigned separate prefixes by the hospital.

### Data Analysis

Continuous variables were reported as median with interquartile ranges (IQR). Categorical variables were expressed as frequencies and percentages. Differences in the distribution of characteristics between the migrant and resident populations were assessed using the Mann-Whitney U test for continuous variables, and Pearson’s chi-square test for categorical variables.

Association between residential status and binary outcomes were examined using logistic regression for rare outcomes with results presented as odds ratios (ORs) and 95% confidence intervals (CIs). These outcomes include: (1) significant injuries (Tier 1 or 2), (2) in-hospital mortality during hospital encounter, and (3) discharge Against Medical Advice (AMA) from the Emergency Department (ED). For ED discharge which is a more common outcome, modified Poisson regression with robust standard errors was applied to directly estimate prevalence ratio (PRs) and 95% CIs, thereby providing a more interpretable measure of association. Patients aged 60 years above were excluded from all regression analyses because unskilled and semi-skilled migrants in Singapore are only permitted to work until that age. For models of AMA discharge and ED discharge, only Tier 3 patients were included, as no Tier 1 or 2 patients were discharged directly from the ED. For model on AMA discharge, the study population used had excluded patients with a direct discharged from ED because these patients do not need to AMA discharge. All regression models were adjusted for age (in 10-year age bands) and sex; in addition, the mortality model was further adjusted for injury severity.

All tests were two-sided, with p-values < 0.05 interpreted as statistically significant. Analyses were performed using R software version 4.1.1 (R Core Team, 2021).

### Ethics Approval

The ethics approval for this study was obtained from the institution’s ethics review board (Singapore Ethics and Compliance Online System (ECOS) reference number 2024/00115).

## Results

### General Demographics and Trends

A total of 313,402 patients were included in the registry during the 10-year period. After excluding 31,555 with missing information on their residential status, gender, or age, the remaining 281,847 were analysed. Of these, 66,719 (23.7%) were migrants.

Compared with residents, migrants tended to be younger and male. Migrants and residents had median ages of 32 (IQR 27-38) and 56 (IQR 37-72) respectively. Males made up 52,124 (78.14%) of migrants and 114,544 (54.24%) of residents. The ethnic distribution in the residents reflects that of the national proportions.^19^ In migrants, the two largest ethnicities were “Chinese” and “Indian”. In addition, there were 43 other listed ethnicities and nationalities, reflecting the cosmopolitan milieu of the migrant population. The annual injury numbers showed a modest decline from 2017 – 2019 in both populations. The large decline in 2020 coincided with COVID-19 lockdown measures. Thereafter, the numbers increased in 2021 and 2022 but did not reach pre-pandemic levels. Table 1 provides the baseline characteristics and trend of injury numbers across the years. (Graphical representation of number trends presented in the Supplementary Materials).

**Table 1.** Baseline characteristics. Values are count with percentages in parentheses unless otherwise stated.

| Characteristic | Migrants (n = 66,719) | Residents (n = 215,128) |
| --- | --- | --- |
| **Gender** |  |  |
| Male | 52,134 (78.14%) | 114,544 (53.24%) |
| Female | 14,585 (21.86%) | 100,584 (46.76%) |
| **Age (years)** |  |  |
| Median (IQR) | 32 (27–38) | 56 (37–72) |
| 21–30 | 29,660 (44.46%) | 36,180 (16.82%) |
| 31–40 | 23,666 (35.47%) | 26,883 (12.50%) |
| 41–50 | 9,952 (14.92%) | 25,892 (12.04%) |
| 51–60 | 1,997 (2.99%) | 33,585 (15.61%) |
| 60–70 | 650 (0.97%) | 34,008 (15.81%) |
| 71–80 | 486 (0.73%) | 30,338 (14.10%) |
| > 80 | 308 (0.46%) | 28,242 (13.13%) |
| **Ethnicity** |  |  |
| Chinese | 15,901 (23.98%) | 155,888 (72.48%) |
| Indian | 15,846 (23.90%) | 22,879 (10.64%) |
| Malay | 955 (1.44%) | 22,050 (10.25%) |
| Caucasian / Eurasian | 146 (0.22%) | 998 (0.46%) |
| Others | 33,448 (50.45%) | 13,261 (6.17%) |
| **Injury Severity (ISS)** |  |  |
| Tier 1 (> 16) | 445 (0.67%) | 3,514 (1.63%) |
| Tier 2 (9–15) | 1,017 (1.52%) | 12,269 (5.70%) |
| Tier 3 (< 9) | 65,257 (97.81%) | 199,345 (92.66%) |
| **Injury Mechanism** |  |  |
| Fall | 17,826 (26.72%) | 105,564 (49.07%) |
| Tools / Objects / Machinery | 18,789 (28.16%) | 18,442 (8.57%) |
| Vehicular accident | 6,065 (9.09%) | 25,676 (11.94%) |
| Interpersonal violence | 3,524 (5.28%) | 9,519 (4.42%) |
| Foreign bodies | 5,774 (8.65%) | 6,347 (2.95%) |
| Sports | 1,572 (2.36%) | 7,229 (3.36%) |
| Bites / stings | 825 (1.24%) | 1,890 (0.88%) |
| Falling object | 155 (0.23%) | 109 (0.05%) |
| Drowning | 4 (0.01%) | 28 (0.01%) |
| Others | 11,299 (16.94%) | 34,194 (15.89%) |
| Unknown | 886 (1.33%) | 6,130 (2.85%) |
| **Patient Disposition from ED** |  |  |
| Discharged | 58,073 (87.04%) | 138,819 (64.53%) |
| AMA discharge<sup>1</sup> | 1,252 (1.88%) | 4,029 (1.87%) |
| Admitted | 6,083 (9.12%) | 65,183 (30.30%) |
| Admitted to high-acuity wards | 638 (0.96%) | 3,210 (1.49%) |
| Transferred to other hospital | 75 (0.11%) | 561 (0.26%) |
| Morgue | 40 (0.06%) | 289 (0.13%) |
| Unknown | 558 (0.84%) | 3,037 (1.41%) |
| **Mortality** |  |  |
| Death during healthcare encounter | 84 (0.13%) | 1,122 (0.52%) |
| **Year of Presentation** |  |  |
| 2013 | 8,637 (12.95%) | 24,424 (11.35%) |
| 2014 | 8,571 (12.85%) | 24,289 (11.29%) |
| 2015 | 8,468 (12.69%) | 25,308 (11.76%) |
| 2016 | 8,082 (12.11%) | 24,838 (11.55%) |
| 2017 | 7,194 (10.78%) | 25,915 (12.05%) |
| 2018 | 6,514 (9.76%) | 24,199 (11.25%) |
| 2019 | 6,288 (9.42%) | 21,555 (10.02%) |
| 2020 | 3,717 (5.57%) | 13,313 (6.19%) |
| 2021 | 4,249 (6.37%) | 14,161 (6.58%) |
| 2022 | 4,999 (7.49%) | 17,126 (7.96%) |

### Disparity in outcomes and mechanisms of injury analysis

Migrants had lower proportions of significant injuries and mortality, but higher proportions of getting AMA discharged and discharged from the ED (Figures 1a-d). After adjusting for age, gender, and additionally for injury severity in the in-hospital mortality model, this pattern persisted: migrant demonstrated lower odds of significant injuries and in-hospital mortality, higher odds of AMD discharge and higher prevalence of ED discharge compared with residents (Table 2. Detailed regression output for all models is presented in the Supplementary Materials).

**Table 2.** Association between residential status and outcome variables.

| Outcome | Crude Odds Ratio (95% CI) | p-value | Adjusted Odds Ratio (95% CI) | p-value |
| --- | --- | --- | --- | --- |
| Significant injuries | 0.702 (0.656 – 0.750) | <0.001 | **0.869 (0.807–0.934)** | **<0.001** |
| Mortality during hospital encounter | 0.534 (0.406 – 0.692) | <0.001 | **0.637 (0.476-0.844)** | **0.002** |
| AMA discharged from ED (Tier 3 only) | 1.605 (1.486 – 1.733) | <0.001 | **1.424 (1.312–1.546)** | **<0.001** |
| Outcome | Prevalence Ratio (95% CI) | p-value | Adjusted Prevalence Ratio (95% CI) | p-value |
| Discharged from ED (Tier 3 only) | 1.066 (1.063 – 1.070) | <0.001 | **1.044 (1.041 – 1.048)** | **<0.001** |

Note:
1. When adjusting for age group, “21–30 years” was taken as reference. All regression models were adjusted for age (in 10-year age bands) and sex; in addition, the mortality model was further adjusted for injury severity. 2. AMA discharge and discharge from ED models included only Tier 3 patients because Tiers 1 and 2 patients were not found to have AMA discharged from ED or discharged from ED. This “effectively” adjusted for injury severity because only mild injuries were analysed. Patients with unknown ED disposition or disposition to the morgue were also excluded. 3. For AMA discharge, the denominator further excluded “Discharged from ED” because it does not make up the at risk population for “AMA discharged from ED”.

**Figure 1a.**
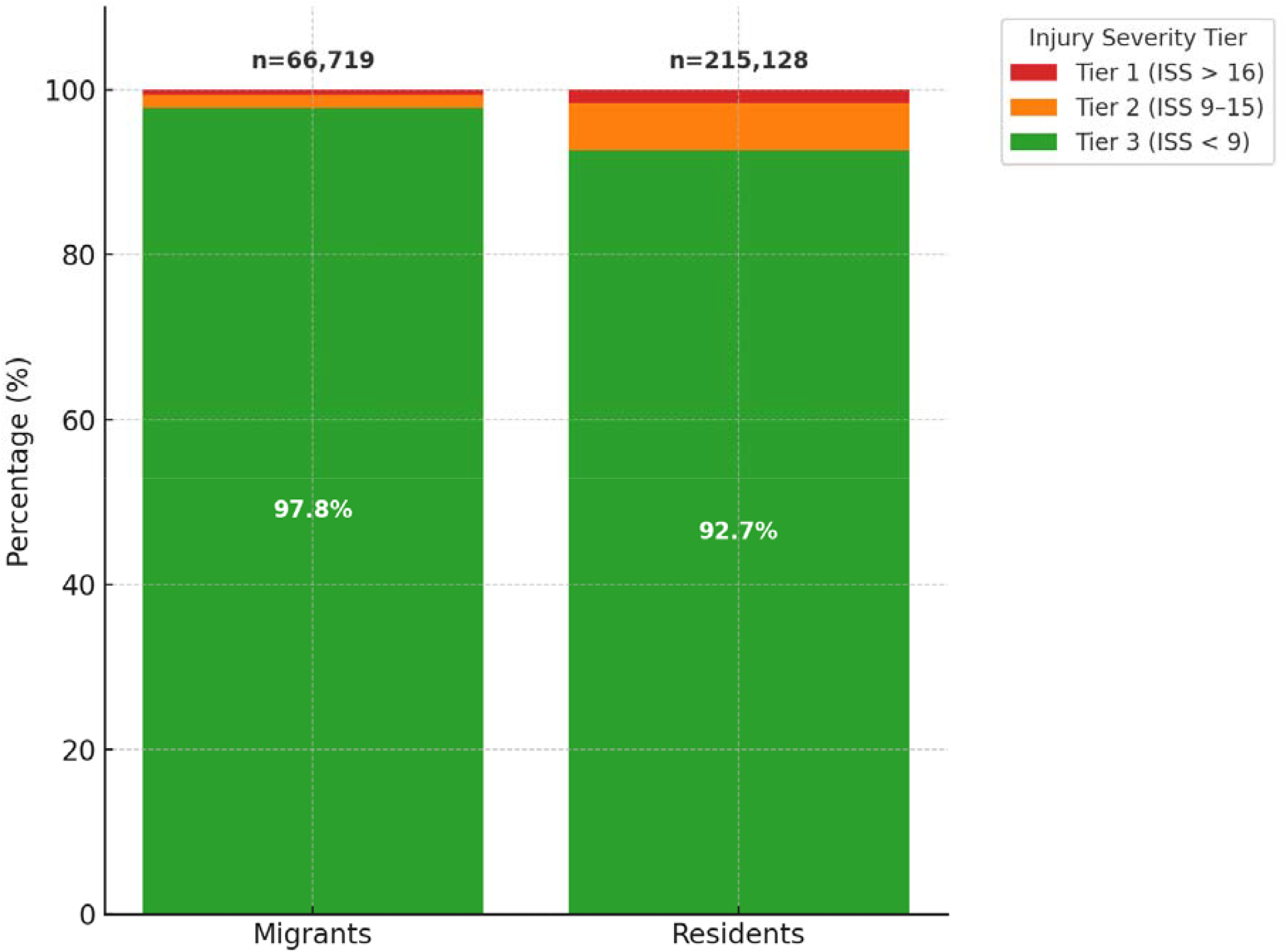
Proportions of significant injuries (Tiers 1 and 2) in migrants and residents.

**Figure 1b.**
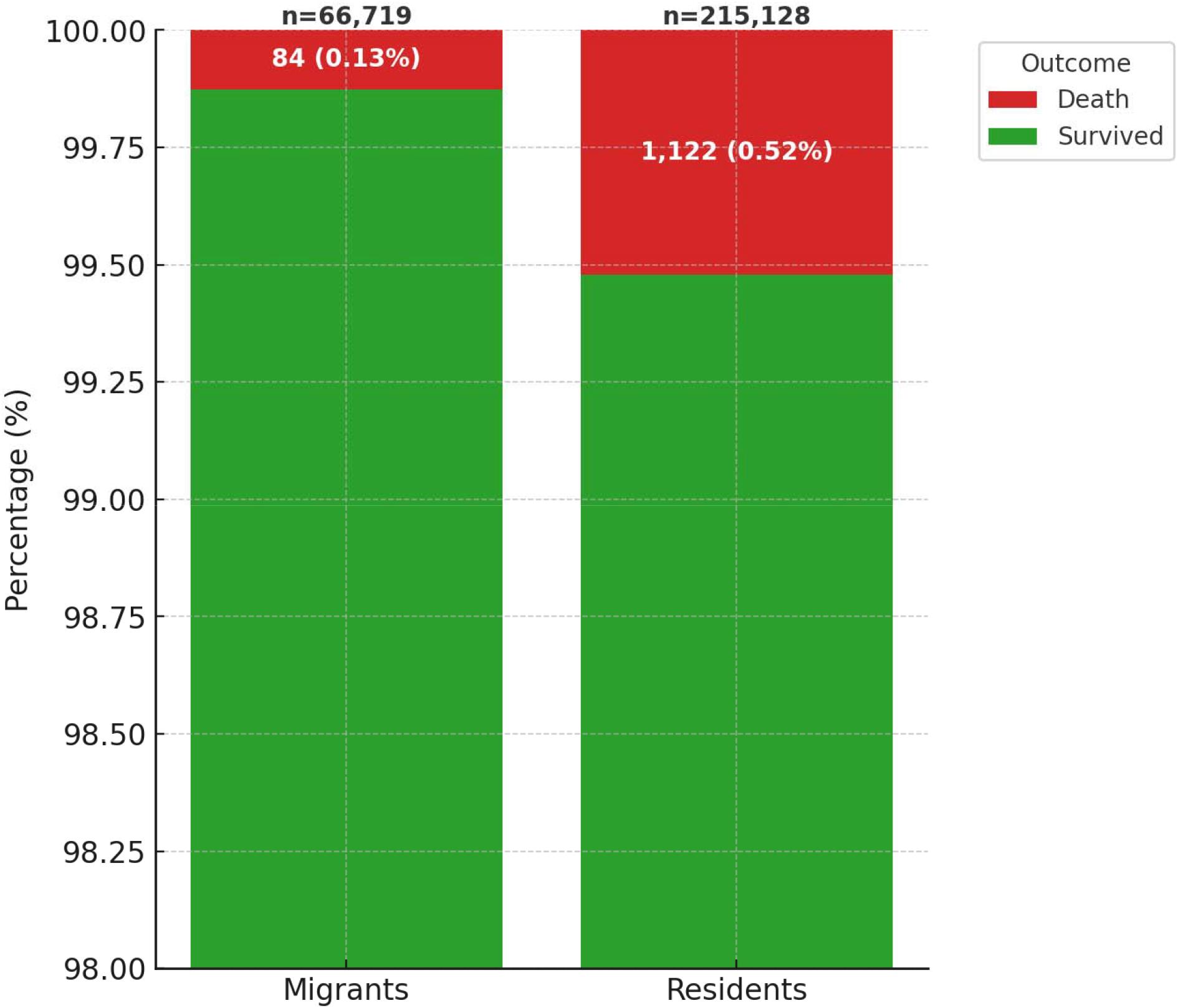
Proportions of mortality during hospital encounter. Y-axis adjusted to highlight the percentage differences.

**Figure 1c.**
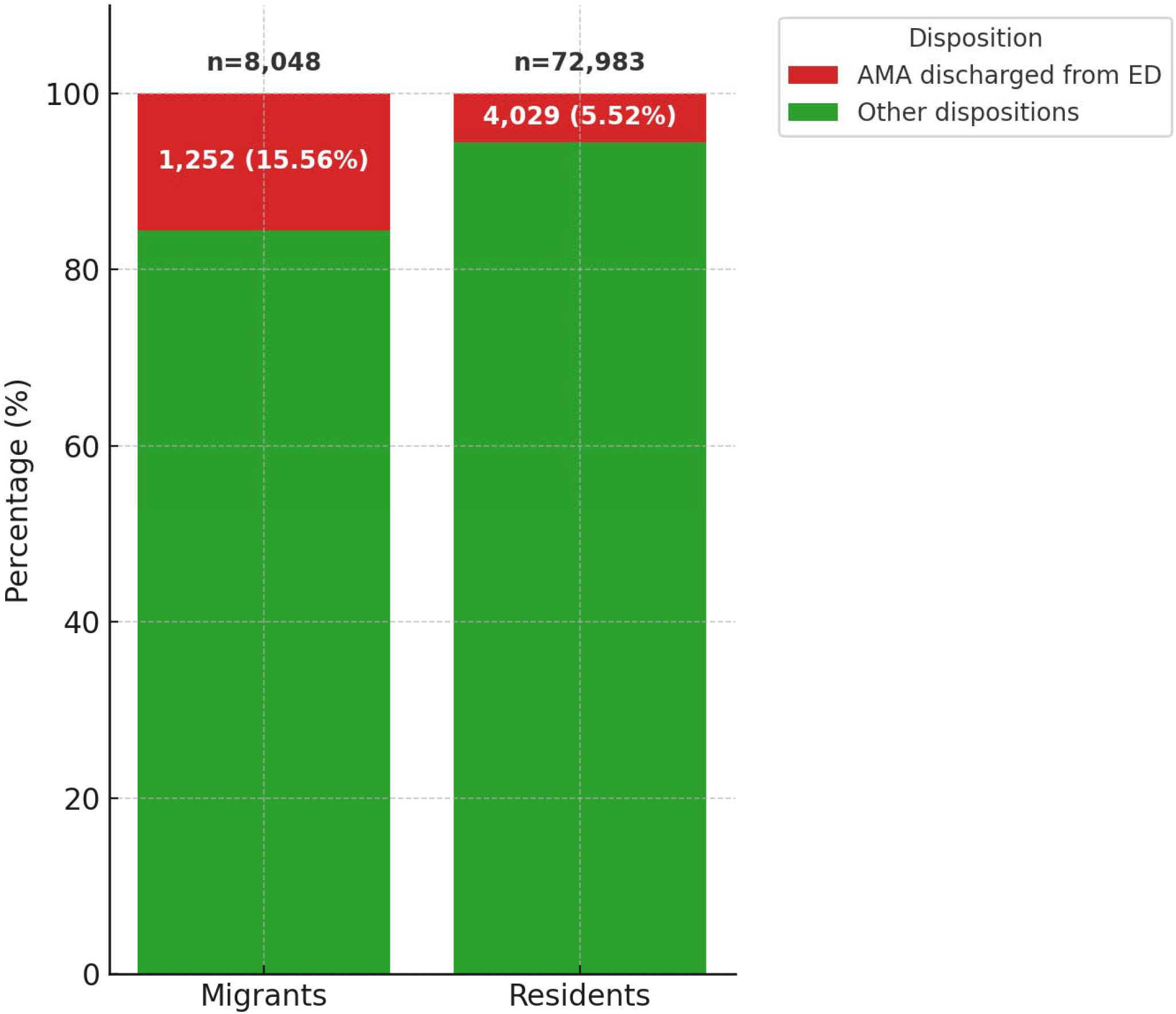
Proportions of AMA discharged from ED.

**Figure 1d.**
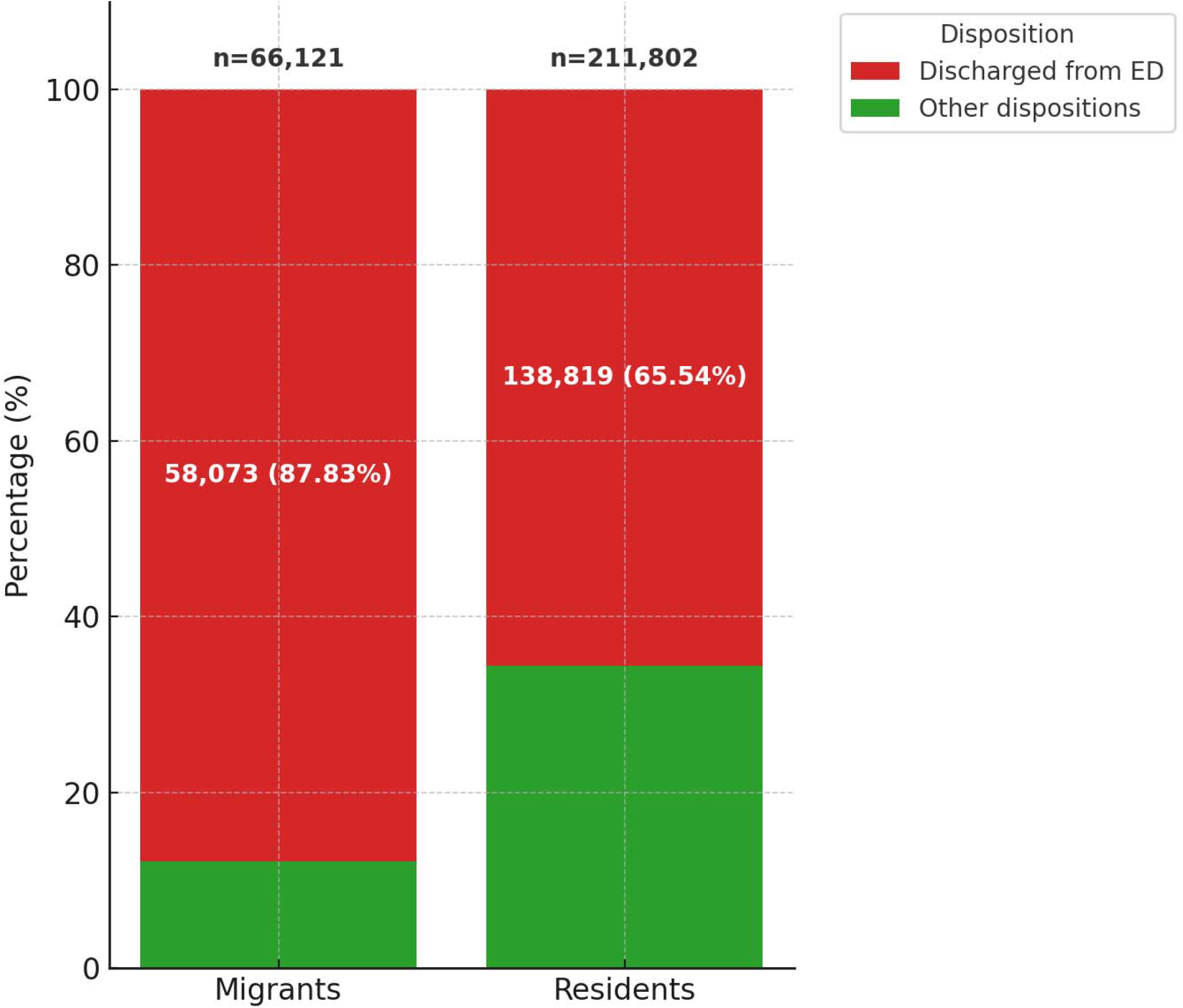
Proportions of “Discharged from ED”.

When all injuries were analysed, the top three injury mechanisms were falls, tools/objects/machinery, and vehicular accidents. However, they were ranked differently. Among migrants, the most common mechanism was tools/objects/machinery, followed by falls, and vehicular accidents. Among residents, the most common mechanism was falls, followed by vehicular accidents, and tools/objects/machinery.

When analysing only significant injuries (ISS ≥ 9), falls became the leading cause of injuries for both migrants and residents. However, migrants tend to fall from height, 50,053 (74.9%), compared with residents 121,036 (53.6%). Vehicular accidents were elevated to rank second in migrants and formed a larger proportion of significant injuries (from 9.1% to 35.8%). Tools/Objects/Machinery dropped from first to third rank in migrants, but in residents, Interpersonal Violence was elevated to third rank, displacing Tools/Objects/Machinery to the fourth rank (Tables 3a, 3b, and 3c).

**Table 3a.**
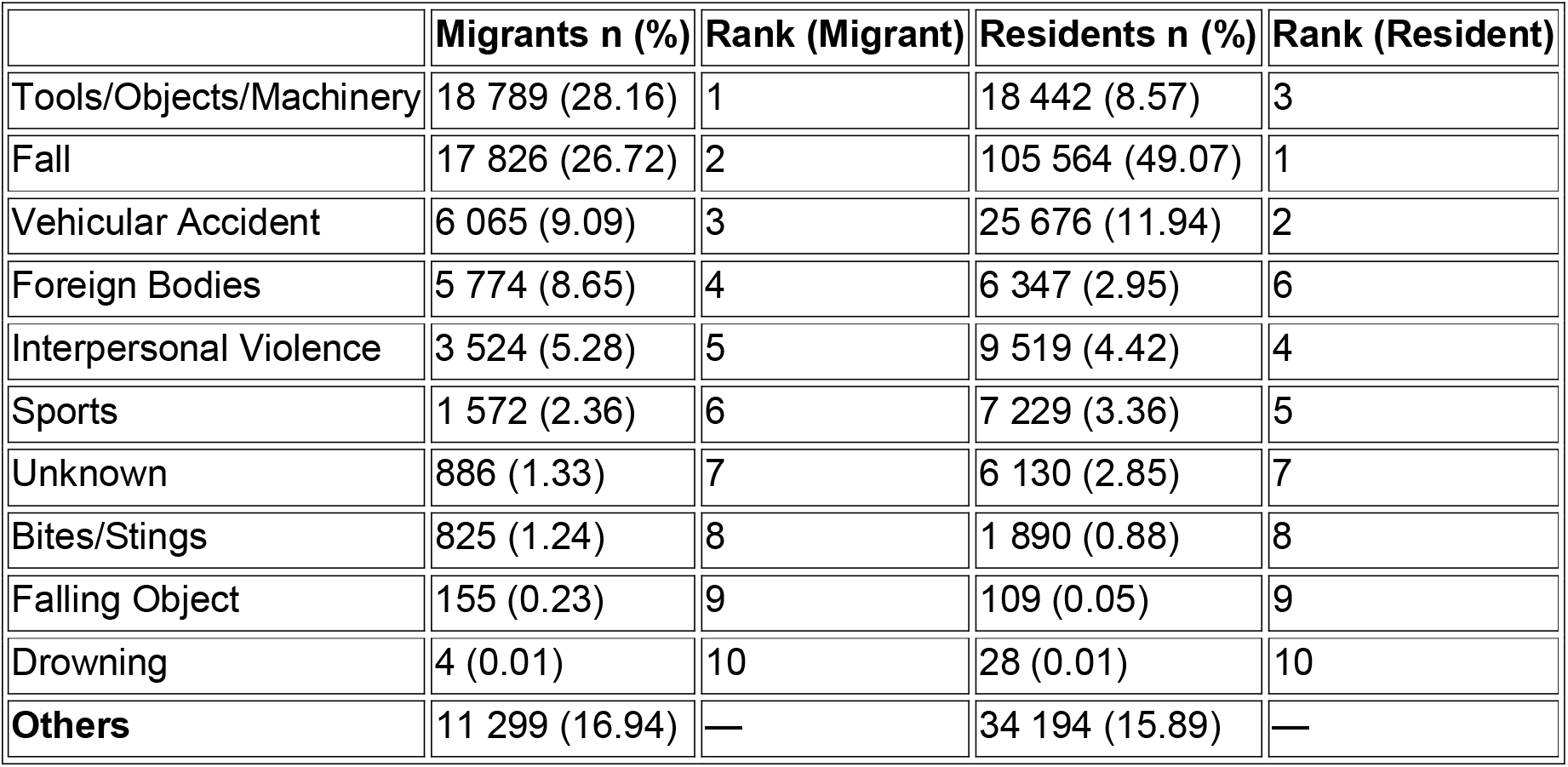
Mechanism of injury among migrants and residents (all injuries)

**Table 3b.** Mechanism of injury among migrants and residents (significant injuries only).

|  | **Migrants (%)** | **Rank (Migrant)** | **Residents (%)** | **Rank (Resident)** |
| --- | --- | --- | --- | --- |
| Fall | 671 (45.89) | 1 | 13,045 (82.72) | 1 |
| Vehicular Accident | 524 (35.83) | 2 | 2,076 (13.15) | 2 |
| Tools/Objects/Machinery | 107 (7.32) | 3 | 147 (0.93) | 4 |
| Interpersonal Violence | 46 (3.15) | 4 | 249 (1.58) | 3 |
| Falling Object | 72 (4.92) | 5 | 25 (0.16) | 7 |
| Foreign Bodies | 14 (0.96) | 6 | 81 (0.51) | 5 |
| Sports | 14 (0.96) | 6 | 68 (0.43) | 6 |
| Drowning | 4 (0.27) | 8 | 28 (0.18) | 8 |
| Bites/Stings | 1 (0.07) | 9 | 2 (0.01) | 10 |
| Unknown | 1 (0.07) | 9 | 16 (0.10) | 9 |
| Others | 8 (0.55) | — | 46 (0.29) | — |

**Table 3c.** Falls by Type.

|  | **Migrants** | **Residents** |
| --- | --- | --- |
| Fall from Height (%) | 50,053 (74.92%) | 121,036 (53.57%) |
| Ground Level Fall (%) | 16,657 (24.93%) | 104,007 (46.04%) |
| Unknown (%) | 9 (0.01%) | 65 (0.03%) |
| **Total** | **66,719** | **225,108** |

## Discussion

This is the largest original research that directly compared injuries and outcomes among migrants and residents from the same catchment.

The proportion of migrants in this study, 23.7%, is lower than the national proportion of migrants (29.2%) in 2018.^20^. The lower proportion could be caused by uneven distribution of injuries to different hospitals. Without the breakdown of the populations at risk in the hospital’s catchment, we could not compare the incidence of injury.

The younger migrant population was likely caused by policies limiting the hiring age for low skilled workers to 60. Injuries are known to affect the working age adult disproportionately.^21^ Our study shows that in a HIC setting, the migrant population contributes significantly to the right skewed age distribution.

The male predominance in migrants reflects the types of industry and work permits issued.^22^ These tend to be jobs in the CMP sector which are deemed risky, onerous, relatively low paying, and shunned by residents, a phenomenon commonly observed globally in HIC settings.^9^

The small decline in injury numbers observed in both populations from 2017 to 2019 could be artefactual given the porous nature of the hospital’s catchment and the opening of a regional hospital in the Northeast of the country in 2019.^23^ However, it is worth noting that both the total migrant and total populations actually increased at the national level during that period.^20,24,25^ Additionally, the decline was preceded by a few workplace safety policies enactment, such as the Work at Heights regulations in 2013,^26^ and the Design for Safety regulations in 2016.^27^ While causal links are speculative, the findings prompt further investigation on the role of legislation on injury prevention.

The better outcome in migrant injury severity and mortality is surprising but may be explained by biases. Firstly, the healthy worker effect suggests that migrants being subjected to pre-employment health checks tend to be healthier than residents.^28^ Another explanation is that migrants with minor injuries preferentially seek help at the hospital instead of primary care facilities.^29^ This selection bias may have inflated the migrant denominator, causing a corresponding decrease in the proportion of significant injuries and mortality. However, assuming that there was a true disparity that migrants suffer poorer outcomes, the above biases would serve to reduce the effect estimate, bringing it closer to the null, resulting a finding of no difference. Yet, our study found a statistical difference in favour of migrants. This is at least a hypothesis generating finding to direct future research, to investigate the protective mechanisms of being in a HIC setting where it is not uncommon for authorities to monitor injury statistics.^30^ In Singapore, the annual publication of workplace safety and health reports by the Ministry of Manpower offered transparency in workplace injuries and mortality statistics.^31^ There was also the Safety Time Out initiative to encourage timely reviews of workplace safety practices.^32^ It is plausible these enacted workplace safety and health measures contributed to the favourable outcomes in migrants.

On barriers to healthcare at the hospital, we found higher rates of discharge from the ED in both AMA discharge and the normal discharge from ED. The decision for AMA discharge is ostensibly made by the patient, suggesting that the hospital is not the cause of the barrier, but the higher adjusted odds suggest that there are other sources of barrier like employer pressure, financial consideration, language difficulties, or cultural differences. In contrast, the normal discharge from the ED decision is made by the physician. This higher proportion of migrant discharge from ED has a few possible explanations which include the healthy worker effect and different health seeking behaviour described earlier.^29^ However, we must also acknowledge the possibility of discrimination and other occult barriers.^1^

The mechanism of injury analysis suggests that amongst migrants, “Tools/ Objects/ Machinery” was a common cause of mild injuries, while “Falls” and “Vehicular Accidents” tend to cause more significant injuries.

Even though falls was the leading cause of significant injuries in both populations, most migrants sustained falls from height, while residents tend to suffer from ground level falls, possibly frailty related. This suggests a greater danger of falling from heights in migrants, but further studies would be needed to establish if these falls were due to work and if further improvement in regulations on working at heights could reduce these falls.

Among cases with significant injuries, the higher proportion of vehicular accidents among migrants was not unexpected given anecdotal observations that migrants and residents commute differently. When analyses were restricted to vehicular accidents, migrants still demonstrated higher odds of sustaining significant injuries compared with residents after adjustment for age and sex (results not shown). There could be a few explanations to this difference. Motorcycles is a popular mode of transport amongst migrants given its affordability.^33^ Singapore is one of few developed countries where the traffic laws allow workers to be ferried at the rear cargo platform of lorries without seatbelts or safety harnesses.^16^ There have been calls to ban such unsafe practices and to transport workers in buses with seatbelts instead, but they were met with resistance by business and trade associations that cited higher operational costs.^34^ Given these findings, future studies may want to look further into vehicular accidents in migrants and residents and investigate the circumstances leading to these accidents.

## Limitations

The descriptive research does not allow for causative claims. The small decline in injury numbers following enacted workplace safety legislations serves only as an observation to direct future research. Interrupted time series research may demonstrate causal relationships between enacted laws and observed improvements.

The trauma registry had limited information on the occupations, whether the injuries were work-related, and the exact mechanism of vehicular injuries, hindering our ability to perform further stratified analysis. Also, the trauma registry does not have co-morbidity data, hence, the logistic regression was not able to adjust for this as a potential confounder.

This study does not capture death on scene cases. The statistics for on-site death for work incidents, captured by the Ministry of Manpower, showed a stable trend of 0.9 to 1.2 deaths per 100,000 workers in the past 8 years,^35^ but does not stratify into migrant versus resident. Other on-site deaths data from vehicular injuries and falls outside of work are captured by the police and is not easily available for analysis.

The lack of denominator data within the hospital’s catchment limited our ability to calculate population-based incidence of injury or mortality. Rather, the study provides insights only on those who presented to the hospital.

Lastly, the single centre study design has limited generalizability. The large sample size provides high statistical power and precision, and the standardized data collection adds to the confidence of the findings’ internal validity. Moreover, public hospitals in Singapore, being funded by the Ministry of Health, have similar capabilities across the city state,^36^ and the Civil Defence is the sole provider of pre-hospital emergency care services. These suggest that the findings may be applicable nationally, but extrapolation to other HIC settings should be made only with comparative data.^37^

## Conclusion

Our study shows that in a HIC setting where migrants typically undertake jobs shunned by residents, migrants do not necessarily suffer a higher proportion of significant injuries and mortality. However, they have higher adjusted odds of getting AMA discharged from ED, and getting discharged from ED, which should raise awareness about healthcare access inequity. This study also provides evidence that migrants suffer a higher proportion of significant injuries caused by fall from heights and vehicular injuries.

Migrants are a vital part of the workforce and contribute greatly to a country’s economic progress. We hope that the knowledge from this study will shine a spotlight on migrants who get injured in their host countries and encourage global dialogue on migrant health and safety.

## Data Availability

All data produced in the present study are available upon reasonable request to the authors.

## Contributors

All authors contributed to the design and implementation of the study. Juin Bin Lee, Hwee Pin Phua, and Natasha Shi Hui Wee performed the data extraction, tidying, and analysis. Xinyi Lin, Yong Jing Daniel Quek, Raj Menon, Li Tserng Teo, Beth Lau, and Wei-Yen Lim contributed to the literature review and iterative writing process. Hong Yee Lo consolidated and edited the final manuscript. All authors approved the final version of the manuscript.

## Acknowledgements

The study team would like to thank the TTSH trauma registry Ms Go Tsung Shyen Karen (manager) and Ms Tong Man (coordinator) for their support in this study.

## Patient and public involvement statement

Patients and members of the public were not involved in the design, conduct, reporting, or dissemination of this study. The analysis was based on de-identified data from a hospital trauma registry, and therefore direct involvement was not feasible. The research question was developed in response to observed disparities in migrant health, and the findings are intended to inform public health policy and healthcare planning to reduce inequities.

## References

1. Lebano A, Hamed S, Bradby H, et al. Migrants’ and refugees’ health status and healthcare in Europe: a scoping literature review. BMC Public Health. Jun 30 2020;20(1):1039. doi:10.1186/s12889-020-08749-8

2. Migrants’ health and persisting barriers to accessing health-care systems. EClinicalMedicine. Feb 2022;44:101321. doi:10.1016/j.eclinm.2022.101321

3. Global research agenda on health, migration and displacement: strengthening research and translating research priorities into policy and practice. 2023. World Health Organisation. Accessed 21 Aug 2025. https://www.who.int/publications/i/item/9789240082397

4. Hargreaves S, Rustage K, Nellums LB, Friedland JS, Zimmerman C, all a. Occupational health outcomes among international migrant workers - Author’s reply. Lancet Glob Health. Dec 2019;7(12):e1616. doi:10.1016/S2214-109X(19)30389-4

5. Quek YJD, Vijayasrinivasan S, Narayanan A, Tham KY. Retrospective review of work-related injuries sustained by foreign workers: a single centre experience over 10 years. BMJ Open. May 10 2021;11(5):e042427. doi:10.1136/bmjopen-2020-042427

6. Ohm E, Holvik K, Kjollesdal MKR, Madsen C. Health care utilisation for treatment of injuries among immigrants in Norway: a nationwide register linkage study. Inj Epidemiol. Nov 16 2020;7(1):60. doi:10.1186/s40621-020-00286-7

7. Ang JW, Chia C, Koh CJ, et al. Healthcare-seeking behaviour, barriers and mental health of nondomestic migrant workers in Singapore. BMJ Glob Health. 2017;2(2):e000213. doi:10.1136/bmjgh-2016-000213

8. Lau K, Aldridge R, Norredam M, et al. Workplace mortality risk and social determinants among migrant workers: a systematic review and meta-analysis. Lancet Public Health. Nov 2024;9(11):e935–e949. doi:10.1016/S2468-2667(24)00226-3

9. Pega F, Govindaraj S, Tran NT. Health service use and health outcomes among international migrant workers compared with non-migrant workers: A systematic review and meta-analysis. PLoS One. 2021;16(6):e0252651. doi:10.1371/journal.pone.0252651

10. Bhattacharjee MR. Mobility and morbidity of regular and seasonal migrants in India. International Journal of Migration, Health and Social Care. 2021;17(2):155–165. doi:10.1108/ijmhsc-04-2020-0038

11. Borhade A, Dey S, Tripathi A, Mavalankar D, Webster P. Migration and health: a review of policies and initiatives in low and middle income countries. The Lancet. 2016;388:S26. doi:10.1016/S0140-6736(16)32262-0

12. Fitzgerald S, Chen X, Qu H, Sheff MG. Occupational injury among migrant workers in China: a systematic review. Injury Prevention. 2013;19(5):348. doi:10.1136/injuryprev-2012-040578

13. Pescarini JM, Goes EF, Pinto Pfps, et al. Mortality among over 6 million internal and international migrants in Brazil: a study using the 100 Million Brazilian Cohort. The Lancet Regional Health – Americas. 2023;20 doi:10.1016/j.lana.2023.100455

14. Population in Brief 2024. Strategy Group, Prime Minister’s Office, Singapore. Accessed 24 Aug 2025. https://www.population.gov.sg/files/media-centre/publications/Population_in_Brief_2024.pdf

15. Foreign workforce numbers. Ministry of Manpower, Singapore. Accessed 2025-03-19. https://www.mom.gov.sg/foreign-workforce-numbers.

16. Dutta MJ. Singapore’s Extreme Neoliberalism and the COVID Outbreak: Culturally Centering Voices of Low-Wage Migrant Workers. Am Behav Sci. Sep 2021;65(10):1302–1322. doi:10.1177/00027642211000409

17. Seah BZQ, Horie S, Gan WH, et al. A comparison of periodic health examinations and workplace health screening for workers in Singapore and Japan. Ind Health. Jan 24 2025;63(1):93–106. doi:10.2486/indhealth.2024-0046

18. aNHG Population Health. Accessed 24 Aug 2025. https://www.nhghealth.com.sg/about-us/nhg-population-health.

19. Frost MR. An unsettled majority: immigration and the racial ‘balance’ in multicultural Singapore. Journal of Ethnic and Migration Studies. 2021/12/10 2021;47(16):3729–3751. doi:10.1080/1369183X.2020.1774112

20. Population in Brief. 2018. Strategy Group, Prime Minister’s Office, Singapore. https://www.strategygroup.gov.sg/images/publicationimages/population-in-brief-2018.pdf

21. Haas B, Jeon S-H, Rotermann M, et al. Association of Severe Trauma With Work and Earnings in a National Cohort in Canada. JAMA surgery. 2020-10-28 2020; doi:10.1001/jamasurg.2020.4599

22. Sector-specific rules for Work Permit. Ministry of Manpower. Singapore. Accessed 24 Aug 2025. https://www.mom.gov.sg/passes-and-permits/work-permit-for-foreign-worker/sector-specific-rules/construction-sector-requirements.

23. SKH Campus Official Opening. Sengkang General Hospital. Singapore. Accessed 30 Aug 2025. https://www.skh.com.sg/archive/skh-campus-official-opening.

24. Population in Brief. 2017. Strategy Group, Prime Minister’s Office, Singapore. https://www.strategygroup.gov.sg/images/publicationimages/population-in-brief-2017.pdf

25. Population in Brief. 2019. Strategy Group, Prime Minister’s Office, Singapore. https://www.strategygroup.gov.sg/files/media-centre/publications/population-in-brief-2019.pdf

26. Workplace Safety and Health (Work at Heights) Regulations 2013 (S 223/2013), Attorney-General’s Chambers, Singapore (2013). Accessed 2025-08-25. https://sso.agc.gov.sg/SL/wsha2006-s223-2013

27. Workplace Safety and Health (Design for Safety) Regulations 2015 (S 428/2015), Attorney-General’s Chambers, Singapore (2015). Accessed 2025-08-25. https://sso.agc.gov.sg/SL/WSHA2006-S428-2015

28. Shah D. Healthy worker effect phenomenon. Indian Journal of Occupational and Environmental Medicine. 2009-05-01 2009;13:77–79. doi:10.4103/0019-5278.55123

29. Chan JSE, Chia DWJ, Hao Y, Lian SWQ, Chua MT, Ong MEH. Health-seeking behaviour of foreign workers in Singapore: Insights from emergency department visits. Ann Acad Med Singap. Apr 2021;50(4):315–324. doi:10.47102/annals-acadmedsg.2020484

30. Selveindran S, Tango T, Khan M, et al. Mapping global evidence on strategies and interventions in neurotrauma and road traffic collisions prevention: a scoping review. Systematic Reviews. 2020-01-02 2020;9 doi:10.1186/s13643-020-01348-z

31. Workplace Safety and Health Report: National Statistics. 2024. Ministry of Manpower, Singapore. https://www.mom.gov.sg/-/media/mom/documents/safety-health/reports-stats/wsh-national-statistics/wsh-national-stats-2024.pdf

32. MOM, WSHC Industry Associations and NTUC Make Joint Call for Safety Time-Out at Workplaces. Accessed 2022-05-09. https://www.tal.sg/wshc/media/press-releases/2022/mom-wshc-industry-associations-and-ntuc-make-joint-call-for-safety-time-out-at-workplaces.

33. Operation Sunrise at Tuas Checkpoint. Accessed 2024/08/28. https://www.ica.gov.sg/news-and-publications/newsroom/media-release/operation-sunrise-at-tuas-checkpoint?utm_source=chatgpt.com. Media Release: Singapore.

34. Zalizan, T. The Big Read: Transporting migrant workers on lorries - why can’t we stop the unsafe practice after so long? CNA Today. Accessed 24 Aug 2025. https://www.channelnewsasia.com/today/big-read/big-read-transporting-migrant-workers-lorries-3709986

35. WORKPLACE SAFETY & HEALTH REPORT, 2024.; 2024. Ministry of Manpower. Singapore. Accessed 6 Sep 2025. https://www.mom.gov.sg/-/media/mom/documents/safety-health/reports-stats/wsh-national-statistics/wsh-national-stats-2024.pdf.

36. Tan CC, Lam CSP, Matchar DB, Zee YK, Wong JEL. Singapore’s health-care system: key features, challenges, and shifts. The Lancet (British edition). 2021;398(10305):1091–1104. doi:10.1016/S0140-6736(21)00252-X

37. Choi SJ, Oh MY, Kim NR, Jung YJ, Ro YS, Shin SD. Comparison of trauma care systems in Asian countries: A systematic literature review. Emerg Med Australas. Dec 2017;29(6):697–711. doi:10.1111/1742-6723.12840

