## Supplemental table 1 and figure 1 for "A 10-Year Comparison of Injury Patterns and Outcomes between Migrants and Residents in a high-income country setting"

Supplementary table 1. Injury severity Analysis (Logistic model of odds of having Tier 1 or 2 severity)

*Excluded individuals older than 60 as very few migrants were above 60.*

|  | **Odds Ratio Estimate** | **Confidence Interval of Odds Ratio** | **P-value** |
| --- | --- | --- | --- |
| **Residential Status** (ref: Residents) |  |  |  |
| Migrants | 0.869 | 0.807 - 0.934 | <0.001 |
| **Gender** (ref: Male) |  |  |  |
| Female | 0.509 | 0.473 - 0.547 | <0.001 |
| **Age Group**  (ref: 21 – 30) |  |  |  |
| 31 – 40 | 1.257 | 1.15 - 1.374 | <0.001 |
| 41 – 50 | 1.715 | 1.566 - 1.878 | <0.001 |
| 51 – 60 | 3.028 | 2.781 - 3.298 | <0.001 |

Supplementary table 2. Analysis of death cases (Logistic model of odds of mortality)

*Excluded individuals older than 60 as very few migrants were above 60. Death was based on death during hospital encounter.*

|  | **Odds Ratio Estimate** | **Confidence Interval of Odds Ratio** | **P-value** |
| --- | --- | --- | --- |
| **Residential Status**  (ref: Residents) |  |  |  |
| Migrants | 0.637 | 0.476 - 0.844 | 0.002 |
| **Gender** (ref: Male) |  |  |  |
| Female | 0.489 | 0.372 - 0.636 | <0.001 |
| **Age Group**  (ref: 21 – 30) |  |  |  |
| 31 – 40 | 1.278 | 0.91 - 1.792 | 0.155 |
| 41 – 50 | 1.833 | 1.312 - 2.564 | <0.001 |
| 51 – 60 | 2.785 | 2.037 - 3.835 | <0.001 |

Supplementary table 3. *AMA Discharge Analysis (Tier 3 only)*

*Excluded individuals older than 60 as very few migrants were above 60 and excluded individuals that ED Disposition was Discharged from ED, Unknown or Morgue.*

|  | **Odds Ratio Estimate** | **Confidence Interval of Odds Ratio** | **P-value** |
| --- | --- | --- | --- |
| **Residential Status** (ref: Residents) |  |  |  |
| Migrants | 1.424 | 1.312 - 1.546 | < 0.001 |
| **Gender** (ref: Male) |  |  |  |
| Female | 0.978 | 0.904 - 1.057 | 0.575 |
| **Age Group**  (ref: 21 – 30) |  |  |  |
| 31 – 40 | 0.908 | 0.825 - 0.999 | 0.049 |
| 41 – 50 | 0.895 | 0.808 - 0.990 | 0.031 |
| 51 – 60 | 0.635 | 0.571 - 0.706 | < 0.001 |

Supplementary table 4. *Discharge Analysis (Tier 3 only).*

*Excluded individuals older than 60 as very few migrants were above 60 and excluded individuals that ED Disposition was Unknown or Morgue.*

|  | **Odds Ratio Estimate** | **Confidence Interval of Odds Ratio** | **P-value** |
| --- | --- | --- | --- |
| **Residential Status** (ref: Residents) |  |  |  |
| Migrants | 1.044 | 1.040 - 1.048 | < 0.001 |
| **Gender** (ref: Male) |  |  |  |
| Female | 1.020 | 1.016 - 1.024 | < 0.001 |
| **Age Group**  (ref: 21 – 30) |  |  |  |
| 31 – 40 | 0.984 | 0.980 - 0.988 | < 0.001 |
| 41 – 50 | 0.953 | 0.948 - 0.958 | < 0.001 |
| 51 – 60 | 0.902 | 0.897 - 0.908 | < 0.001 |

Supplementary figure 1.

Injury number trend across the 10-year study period from 2013 – 2022, stratified by residential status.

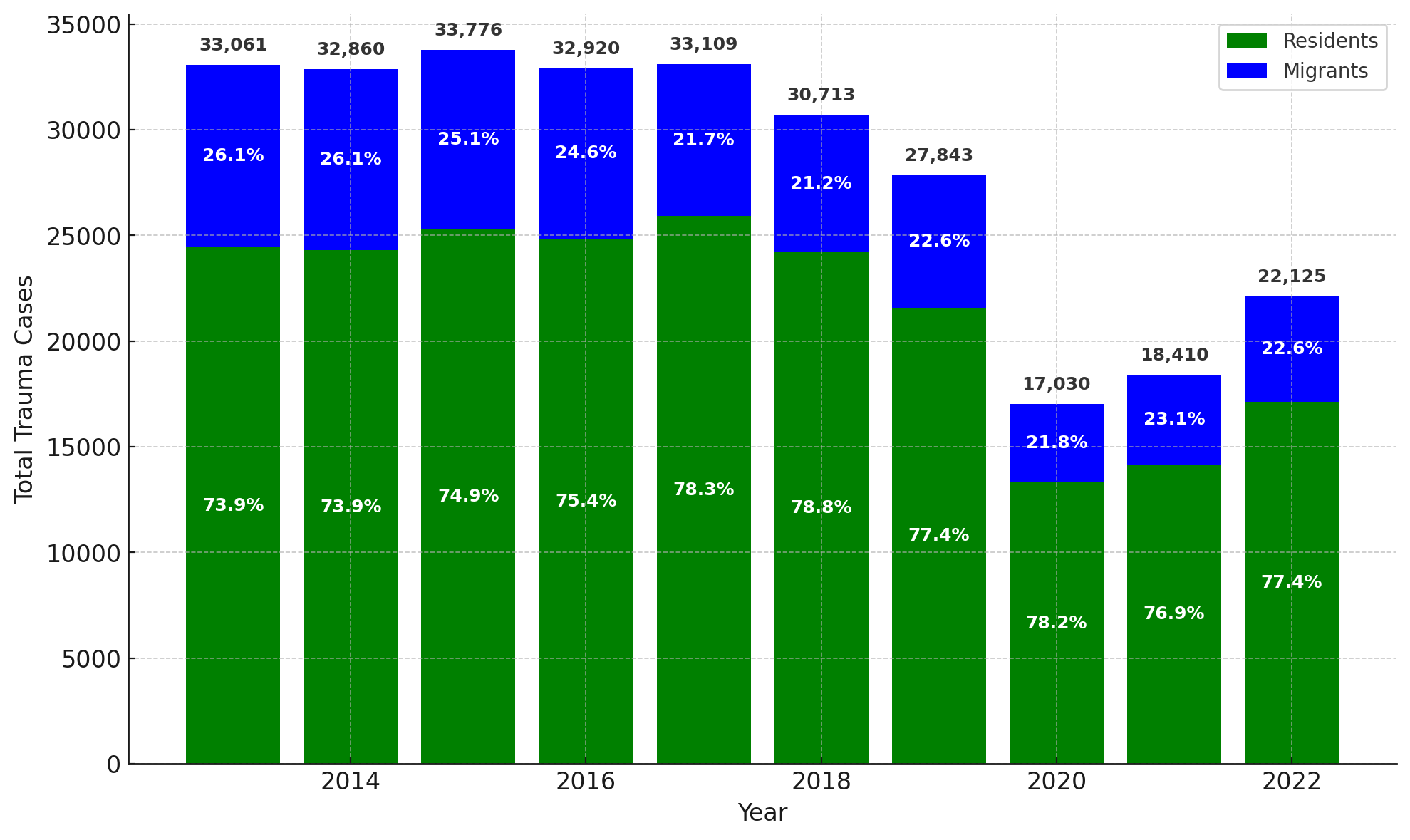
